# Assessing the effectiveness of animal therapy on alleviation of anxiety in pre-school children

**DOI:** 10.1101/2020.08.30.20184986

**Authors:** Mohammad Tahan, Azam Sadeghifar, Elahe Ahangri

## Abstract

**Introduction:** The purpose of this study was to assess the effectiveness of animal therapy in alleviation of anxiety in pre-school children.

**Method:** The study was carried out as a quasi-experimental study with pre-test and post-test design and control group. The study population consisted of 33 anxious 5-7years old children (participated in a welfare anxiety screening plan held by Counseling Center, Tehran-Iran) between 2018 and 2019. The participants took part in the study voluntarily.

The subjects were randomly divided into experimental and control groups (10 in each group). The experimental group was exposed to 8 sessions of animal therapy. The research instrument was Spence Preschool Anxiety Scale (Parent Form) and the data were analyzed on SPSS 21 software.

**Results:** The results showed that animal therapy had a significant effect on general anxiety score after adjusting for post-test scores (f= 32.49 and p= 0.001) with the effect equal to 0.70. In addition, the effect of animal therapy on anxiety of separation (f= 5.63, p= 0.03), generalized anxiety disorder (f= 8.56, p= 0.01), social phobia (f= 14.58, p= 0.002) and specific anxiety (f= 11.63, p= 0.005) was significant with effects equal to 0.30, 0.40, 0.53, and 0.47, respectively. The results also showed that the effect of animal therapy on obsession was not significant (p>0.05).

**Conclusion:** Therefore, it can be concluded that Animal therapy is effective in alleviating anxiety in children.

## Introduction

Children constitute a large portion of the world population. Their physical, emotional, mental, and behavioral growth have been always a concern of scientists and researchers. Over the past 25 years, behavioral, social, and emotional problems of children have been among the main and critical issues in psychology and psychiatry (Mash & Barkley, 2014).

Epidemiology studies have shown that with a prevalence range of 5-17%, anxiety is one of the main psychological disorders in childhood (Castlo and Angold 1995; cited by Haidari et al., 2016). Anxiety disorder is one of the common disorders in childhood and it can lead to other disorders. Such disorder intervenes with function in other fields and usually appears along with other disorders like depression and uncontrollable behavioral disorders. Children with anxiety disorder are at a high risk of committing suicide and psychology disorders when they become teenagers or adults (Matin, 2009).

By anxiety, we refer to unpleasant emotions that are expressed by terms like “worries,” “fear,” and “panic.” Everyone experiences different levels of anxiety during their life (Atkinson, et al. 2016). It is an indispensable part of childhood and a sign of normal growth of children. In fact, it may have positive effects on child’s growth as it gives the child a chance to develop coping strategy and to deal with stressors and causes of anxiety in the future (Hughes, 1996). Anxiety, in its comparative form, helps children to adapt to others’ world. A normal level of anxiety has a regulating function and helps the child to adapt their behaviors to social, educational, and cultural expectations. On the other hand, too low and too high anxiety can be a cause of maladaptation. The individuals who demonstrate antisocial behaviors or behaviors related to conduct disorder are usually do not have anxiety arousal. Constant and excessive anxiety also causes maladaptation, which leads to distress and interruption of development process (Graham, 2003).

According to Silverman (2011), although, the experience of fears and worries, as a part of the development process, is temporary in many children, some of them have this experience for a longer period of time and with high intensity so that it disrupts their daily functions (Mychailyzvn, Mendez, and Kendall 2010). According to Diagnostics and Statistical Manual of Mental Disorders (DSM-V) anxiety disorders diagnosed in children are separation anxiety disorder, opted muteness, specific phobia, social anxiety, panic anxiety, generalized anxiety, anxiety caused by drugs, and anxiety caused by physical disease. A common aspect in all these disorders is that they appear as special and non-continuous responses, cognitive organs responses (physiological), and behavioral responses (American Psychiatric Association, 2018).

Anxiety and the resultant disorders degrade abilities of children notably and lead to problems in doing daily activities, inter-personal relationships, social skills, relationships with peers, and educational performance (Barrett, 1998; Dweck and Wortman 1982; Last, hanson and Franco 1997). Anxiety is a risk factor for other disorders and anxiety related disorders in particular (Cole et al., 2000).

Taking into account the prevalence of anxiety disorders in children, several therapeutic and educational programs have been introduced for adaptation to, prevention of, and treatment of them (Haidari et al. 2016). Yoosefi et al. (2008) showed that narrative therapy alleviated anxiety symptoms and comorbid anxieties in children with anxiety. Hudson et al. (2005) showed that cognitive behavioral therapy was effective in anxiety in children. In addition, there has been an increase in attention to animal-assisted therapy (AAT) in the recent years. Researchers believe that AAT can be considered as a supplementary treatment for traumas (Marguerite, Noemie and Alison, 2015). The AAT treatment is not limited to a specific group of disorders (Worsman 2016). Worsman (2016) and Ines Pandzic (2016) highlighted the effects of AAT to treat the traumas after an accident. They showed that this method can be used to alleviate anxiety and that, as companions, animals function as social facilitators.

Throughout history, animals have been used for different therapeutic purposes (Serpell, 2010). In general, AAT is defined as any intervention with participation of an animal as a part of the process (Kruger and Serpell, 2010). By process, we refer to purposeful therapeutic interventions using animals (AAT), less structured enrichment activities with animals (animal-assisted activities), and using trained animals to help in performing daily tasks (services or supporter animals) (Markoverity et al., 2015). The AAT is a sort of treatment aimed at creating a therapeutic intervention for humans using animal in the treatment process. This treatment is focused on improving behavioral, social, emotional, cognitive, and physical performance. In most of the cases, AAT is a structured intervention with specified purposes that leads to measurable and well-defined outcomes that are listed the Int’l association of human animal interaction organization (IAHAIO) (Aleksandrowicz, Avent and Hassiotis, 2016). As social facilitators, animals can communicate with people and act as a reminder of peace (Chumley et al., 2013), or a safe zone (Parish-Plass, 2008). Through this, animals can alleviate the sense of loneliness and isolation and create connections with humans (Banks and Banks, 2002). Markoirit et al. (2015) showed in a review paper that AAT alleviated depression, PTSD, and anxiety. Crump (2015) showed that PT programs led to less stress in people and the literature indicates that exposure to PT may attenuate physiological stress, mental stress, and anxiety level (Crump, 2015). Given this introduction and the absence of studies in Iran on AAT and its effect of anxiety in children, and given the fact that children nowadays are faced with a variety of challenges and problems that affect their anxiety level, it is imperative to fine reliable ways to prevent anxiety in children. Prevention of emotional behavioral problems in children using timely intervention is supported by many studies. Therefore, the present paper tries to answer if AAT is effective on alleviation of anxiety in pre-school children?

## Methodology

The study was carried out as a semi-experimental study with pretest/posttest design and a control group. The study population consisted of all children at the age of 5 to 7 years with anxiety (referred by Tehran Welfare Anxiety Screening Program to Consultation Center; n=32). Totally, 20 children with an anxiety score higher than the average score were selected using convenient sampling. The participants were assigned to a control and experiment groups (n=10). The experiment group received eight AAT sessions and the control group received no intervention. Before and after the intervention, the mothers filled in Spence Children Anxiety Scale for parents (SCAS-P) and a structured interview based on DSM-V (pretest and posttest) was conducted. Inclusion criteria were anxiety score above the average score, no psychological disorder, no physical disorder, tendency to participant in the study, and age range of 5-7 years.

Data gathering tools included SCAS-P, structured interview based on DSM-V, and a demographics form.

### Spence Children Anxiety Scale for parents (SCAS-P)

The scale is designed to measure anxiety based on DSM-IV diagnosis standard by Spence et al. (Rapee, Kennedy and Ingram 2010). There are 28 statements in the tool with five sub-scales of separation anxiety (five statements), generalized anxiety (five statements), social anxiety (six statements), agoraphobia (seven statements), and compulsive obsessive disorder (five statements). The scale is designed for the parents to measure anxiety in children at 2 -6 years old range. The statements are designed based on Likert’s five-point scale (1= never like this, …, 5=always like this). The higher the score the higher the anxiety in the child. Bassak nejad et al. (2012) obtained reliability of the scale using Cronbach alpha equal to 0.89 and validity of the whole tool was obtained at 0.01 and 0.57 level. In addition, test register prediction and total diagnosis power of the test were obtained equal to 0.70 and 0.76 respectively. Among the sub-scales, agoraphobia, social anxiety, and separation anxiety have a higher validity.

### Implementation

After securing a letter or permission from the head of Tehran City Iranian Consultation Center and creating a good relationship with the parents of children with anxiety (referred to the center by Mashhad City Welfare Org.; n = 32), the parents who were interested in participation in the study were asked to sign a written letter of consent for their child’s participation in the study. The participants were ensured that their personal and private information will remain confidential and that the study is in compliance with religious and cultural codes. Afterwards, the parents filled out SCAS-P and participated in a structured interview based on DSM-V (pre-test). Then, 20 children with anxiety score above the average were selected through convenient sampling method and randomly assigned to experiment and control groups (each with 10 members). The experiment group received eight AAT sessions (each 90 min) and afterwards, the participants filled out the questionnaire once more (post-test).

**Table 1.** Content of the therapeutic protocol.

|  |  |
| --- | --- |
| **Session one** | Introduction, briefing, primary assessing and introduction to the course |
| **Session two** | Giving an introduction about animals and their specific characteristics |
| **Session three** | Taking children to the place that animals were kept |
| **Session four** | Spending time in the place that animals were kept, talking about them and how to communicate with them |
| **Session five** | Convincing children to come closer and touch and pet them |
| **Session six** | Playing with animal and feeding them |
| **Session seven** | Participation in activities like washing and feeding the animals |
| **Session eight** | Summarizing, giving feedback, and post-test |

### Findings

Descriptive statistics showed that pretest and posttest mean scores of anxiety in the experiment group were 67.90 and 55.30 respectively-i.e. 12.60 points decrease in anxiety. In contract, pretest and posttest mean scores of anxiety in the control group were 69.20 and 70.0 respectively – i.e. 0.80 point increase. As to the subs-scale anxiety of separation, the pretest and posttest mean scores in the experiment group were 12.60 and 10.40 respectively (2.20points decrease); these figures in the control group were 12.40 and 12.70 respectively (0.30point increase). With regard to generalized anxiety, pretest and posttest mean scores in the experiment group were 12.40 and 9.50 respectively (2.90 points decrease); in the case of control group these figures were 12.90 and 13.10 respectively (0.20point increase). The pretest and posttest mean scores of social anxiety in the experiment group were 13.70 and 10.40 respectively (3.30 points decrease); these figures in the control group were 14.30 and 14.80 respectively (0.50point increase). As to agoraphobia, the pretest and posttest mean scores in the experiment group were 16.80 and 13.20 respectively (3.60points decrease); as to the control group, these figures were 17.30 and 17.10 respectively (0.20point decrease). Finally, at to obsession, the pretest and posttest mean scores in the experiment group were 12.40 and 11.80 respectively (0.60point decrease); these figures in the control group were 12.30 and 12.30 respectively (no change). In addition, ANCOVA was used for data analysis and before that homogeneity of variances and consistency of regression line slop were checked.

**Table 2.** Leven’s test to examine homogeneity of variances.

| Variables | F-value | DF 1 | DF 2 | P-value |
| --- | --- | --- | --- | --- |
| Total score of anxiety | 0.003 | 1 | 18 | 0.957 |
| Anxiety of separation | 0.244 | 1 | 18 | 0.628 |
| Generalized anxiety | 1.247 | 1 | 18 | 0.279 |
| Social phobia | 0.051 | 1 | 18 | 0.824 |
| Special phobia | 0.518 | 1 | 18 | 0.481 |
| Obsession | 0.552 | 1 | 18 | 0.467 |

**Table 3.** Variance analysis to examine homogeneity of variance.

| Variables | Source | Sum of Squares (SS) | DF | F-value | P-value |
| --- | --- | --- | --- | --- | --- |
| Total score of anxiety | Group interaction with pre-test | 8.450 | 1 | 0.166 | 0.689 |
| Anxiety of separation |  | 0.200 | 1 | 0.077 | 0.785 |
| Generalized anxiety |  | 1.250 | 1 | 0.456 | 0.508 |
| Social phobia |  | 1.800 | 1 | 0.489 | 0.493 |
| Special phobia |  | 1.250 | 1 | 0.263 | 0.615 |
| Obsession |  | 0.050 | 1 | 0.028 | 0.870 |

**Table 4.** ANCOVA results.

| Variables | The sum of the squares (SS) | DF | Mean squares (MS) | F-value | P-value | Power |
| --- | --- | --- | --- | --- | --- | --- |
| Total score of anxiety | 893.784 | 1 | 893.784 | 40.254 | 0.001 | 0.703 |
| Anxiety of separation | 21.972 | 1 | 21.972 | 5.628 | 0.034 | 0.302 |
| Generalized anxiety | 33.097 | 1 | 33.097 | 8.556 | 0.012 | 0.397 |
| Social phobia | 56.002 | 1 | 56.002 | 14.583 | 0.002 | 0.529 |
| Special phobia | 40.321 | 1 | 40.321 | 11.634 | 0.005 | 0.472 |
| Obsession | 1.122 | 1 | 1.122 | 0.448 | 0.515 | 0.033 |

**Table 5.** Mean scores of anxiety and the subscales in the two groups before and after the intervention.

| Variables | Mean difference between pre and post scores in intervention group | Mean difference between pre and post scores in control group | Differences between experimental and control groups | t | P |
| --- | --- | --- | --- | --- | --- |
| Total score of anxiety | -12.60 | 0.80 | -13.40 | -6.539 | 0.001 |
| Anxiety of separation | -2.20 | 0.30 | -2.50 | -3.019 | 0.007 |
| Generalized anxiety | -2.90 | 0.20 | -3.10 | -3.845 | 0.001 |
| Social phobia | -3.30 | 0.50 | -3.80 | -4.971 | 0.001 |
| Special phobia | -3.60 | -0.20 | -3.40 | -4.562 | 0.001 |
| Obsession | -0.60 | 0.001 | -.60 | -.836 | 0.414 |

## Discussion and conclusion

Effectiveness of AAT on alleviation of anxiety in pre-school children was examined. The ANCOVA results showed that AAT was effective in the total score of anxiety after adjusting post-test scores (f=32.49; p=0.001) with effect size of 0.70. In addition, the effect on the subscales anxiety of separation (f=5.63; p=0.03), generalized anxiety (f=8.56; p=0.01); agoraphobia (f=15.48; p=0.002), and social phobia (f=11.63; p = 0.005) was significant with effect sizes of 0.30, 0.40, 0.53, and 0.47 respectively. The effect of AAT on obsession was not significant (p>0.05).

The results are consistent with Tournier (2017), Badr and Zauszniewski (2017), Ines Pandzic (2016); and Aleksandrowicz et al. (206). Worsman (2016) showed that AAT was effective in anxiety; so that, animals function as companions and social facilitators.

Anxiety disorders are of the most common types of psychological disorders in children at school age. It is estimated that the prevalence of this anxiety ranges from 4 to 19% (Ford, Goodman and Meltzer 2003; Komijani, 2010). Anxiety disorders usually appear with other disorders like depression and disruptive behavioral disorders. Children with anxiety disorder are at a high risk of schizophrenia, drug abuse, suicide, and hospitalization in mental hospital during adolescence and young ages (Komijani, 2010). Anxiety is a stable disorder and causes dysfunction in areas like educational performance, self-esteem, depression, and dependence (Merikangas & Avenevoli, 2002).

Several studies have shown that childhood anxiety is a serious mental health problem (Visagie, Loxton and Silverman, 2015). Anxiety disorders have a high rate of comorbidity with other psychological disorders in children (Monga, Brimaher & Ciappetta, 2000). Different interventions like behavioral, cognitive behavioral, and medicine therapy are used to treat anxiety disorder. New therapeutic treatments have been also introduced in the recent years, such as AAT, which is a novel treatment for a wide range of diseases. A large volume of literature has showed that interaction between patient and animal prepares a ground to boost self-confidence in the patient, alleviate the symptoms, and improve the quality of life. Short-term contacts or looking after animals in long-run (horse, cat, birds, small animals) alleviate a large number of diseases (Ines Pandzic, 2016). In addition, there might be several reasons to explain the effectiveness of AAT. Theory of attachment is one of the theories that tries to explain this. Accordingly, attachment is a stable and emotional bond between two individuals (Khanjani, 2005; Geist, 2011). In the case of relationship between man and animal, the emotional bond is formed with the animal. There have been several studies on the relationship of man and animal based on the theory of attachment. There are strong supports of the emotional ties and sensitive bonds between domestic animals and their owners (Prato-Previde et al. 2003; Jasperson, 2010).

In fact, pets play the role of an unconditional love supplier in families (Southerland, 2007; McDowell, 2005). In addition, given the easy access to animal and low costs of keeping pets, they can be used as a reliable and effective treatment. Prothmann et al. (2006), surveyed the effects of animal therapy on mental problems and concluded that AAT improve awareness and range of attention. Pets have a lot of benefits for children and adults; they facilitate making positive social interactions with others, improve daily activity of the owner, facilitate personal relationship, improve sympathy, boost self-confidence, create a sense of safety, and improve communicational and social skills in general (Ascione et al., 2007; Barak et al., 2001; Born, 2010). Lack of these skills may cause anxiety and through improving these skills, AAT helps the individual to create relationship with others, improve supportive resources, and alleviate anxiety in return. Through improving daily activities, AAT attenuates anxiety and improves psychological situation of the individual. A higher health condition, less stress, and fewer behavioral and psychological problems all have been reported as the results of AAT (Allen et al., 2002; Itzchak and Zachor, 2010; Prothmann et al., 2006). Therefore, it can be concluded that AAT is effective in alleviating anxiety in children. As to limitation of the present study, the small number of children in the sample group and number of AAT session are notable. Using other domestic animals or doll animals for AAT purposes can be subject of future studies.

## Data Availability

The raw data supporting the conclusions of this manuscript will be made available by the authors, without undue reservation, to any qualified researcher.

## Ethical Consideration

In the present study, the parents who were interested in participation in the study were asked to sign a written letter of consent for their child’s participation in the study. The participants were ensured that their personal and private information will remain confidential and that the study is in compliance with religious and cultural code. This study has been reviewed and approved by the Institutional Review Board of Omid and Neshat Institute of Tehran with the number 0128/1398. this study registered in Chinese Clinical Trial Registry by number ChiCTR2000034145.

## Acknowledgment

The authors thank Mr Tahan, for their work in data collection and project administration, as well as anonymous reviewers.

## Research involving animals

Not applicable .

## Author Contributions

MT designed the study and conducted the literature searches, wrote the first draft of the manuscript. EA and AS revised the draft of the manuscript. All authors approved the final version of the manuscript.

## Consent for publication

Not applicable.

## References

Aleksandrowicz, S. M., Avent, C & Hassiotis, A. (2016). A Systematic Review of Animal-Assisted Therapy on Psychosocial Outcomes in People with Intellectual Disability. Research in Developmental Disabilities, 49–50, 322–338. Doi: 10.1016/j.ridd.2015.12.005

Allen, K., Blascovich, J., & Mendes, W. B. (2002). Cardiovascular reactivity and the presence of pets, friends, and spouses: the truth about cats and dogs. Psychosomatic Medicine, 64, 727–739. Doi: 10.1097/01.PSY.0000024236.11538.41

American Psychiatric Association. (2018). Diagnostic and Statistical Manual of Mental Disorders. Translated by Yahya Seyed Mohammadi. Tehran: Nashr Ravan.

Ascione, F. R., Weber, C. V., Thompson, T. M., Heath, J., Maruyama, M., & Hayashi, K. (2007). Battered pets and domestic violence: Animal abuse reported by women experiencing intimate violence and by nonabused women. Violence Against Women, 13, 354-373. Doi: 10.1177/1077801207299201

Atkinson, Richard., Atkinson, Rita., Edward, Smith., Nolen-Hoxma, Susan. (2016). Hilgard’s Introduction to Psychology. Translated by Reza Zamani, Mehrdad Beek, Behrouz Birashk, Mohammad Taghi Barahani, and Mehrnaz Shahr-Ara. Tehran: Roshd Publiction.

Badr, H. A., Zauszniewski, J. A. (2017). Kangaroo care and postpartum depression: The role of oxytocin. International Journal of Nursing Sciences, 4, 179-183. Doi: 10.1016/j.ijnss.2017.01.001

Banks M. R., Banks W. A. (2002). The effects of animal-assisted therapy on loneliness in an elderly population in long-term care facilities. J. Gerontol. A Biol. Sci. Med. Sci. 57, M428–M432. Doi: 10.1093/gerona/57.7.M428

Barak, Y., Savorai, O., Mavashev, S. & Beni, A. (2001). Animal-assisted therapy for elderly schizophrenic patients: A one-year controlled trial. American Journal of Geriatric Psychiatry, 9, 439-442. Doi: 10.1097/00019442-200111000-00013

Barrett, P. M. (1998). Evaluation of cognitive-behavioral group treatments forchildhood anxiety disorders. Journal of Clinical Child Psychology, 27, 459–468. Doi: 10.1207/s15374424jccp2704_10

Bassak Nejad, S., Poloi Shahpor Abadi, F., Davoudi, I. (2012). Efficacy of Family Anxiety Management Training of Mothers with Anxious Kindergarten Children aged between 4 -6 Years. Jundishapur Scientific Medical Journal, 11(4), 365-373.

Born, A. (2010). The relationship between humans and animals in animalassisted therapy: A qualitative study. US: ProQuest Information & Learning, 71(1-B), 650. ISBN 978-1-109-56395-5

Cole DA, Peeke LA, Martin JM, Truglio R, Seroczynski AD (2000). A longitudinal look at the relation between depression and anxiety in children and adolescents. Journal of Consulting and Clinical Psychology, 106:586–597.

Crump, C., & Derting, T. L. (2015). Effects of pet therapy on the psychological and physiological stress levels of first‐year female undergraduates. North American Journal of Psychology, 17, 575–590. Retrieved from http://najp.us/

Dweck, C. S., & Wortman, C. B. (1982). Learned Helplessness, Anxiety, and Achievement Motivation: Neglected Parallels in Cognitive, Affective, and Coping Responses. In H. W. Krohne, & L. Laux (Eds.), Achievement, Stress, and Anxiety (pp. 93-125). Washington DC: Hemisphere.

Ford, T., Goodman, R., Meltzer, H. (2003). The British Child and Adolescent Mental Health Survey 1999: the prevalence of DSM-IV disorders. J Am Acad Child Adolesc Psychiatry, 42(10):1203-11. Doi: 10.1097/00004583-200310000-00011

Geist, T. S. (2011). Conceptual Framework for Animal Assisted Therapy. Child Adolesc Soc Work J, 28, 243-256. Doi: 10.1007/s10560-011-0231-3

Graham, Philip. (2003). Cognitive behavioral therapy for children and families. Translated by Mohammad Reza Mohammadi, Hooshmand Hashemi Kohanzadeh. Tehran: University of Social Welfare and Rehabilitation Sciences.

Haidari, M., Bakhtiyarpoor, S., Makvandi, B., Naderi, F., hafezi, F. (2016). Investigating the Effectiveness of FRIENDS Program Training on the Anxiety of Children in Shiraz. Journal - Research Methods and psychological models, 7(24), 23-38.

Hudson, J.L., Devencye. And Taylor L. (2005). Nature, Assessment, and Treatment of Generalized Anxiety Disorder in Children. Pediatr Ann, 34(2): 97-106. Doi: 10.3928/0090-4481-20050201-08

Hughes, John. Ann. (1996). Clinical Child Psychology. Translated by: Najariyan, Khodarimi and Makundi. Ahwaz: Mardomak Publishing. Doi: 10.1111/j.1469-7610.1996.tb01396.x

Ines Pandzic, M. A. (2016). Animal-Assisted Therapy and PTSD. http://www.med.navy.mil/sites/nmcsd/nccosc/healthProfessionalsV2/reports/Documents/white-paper-animal-assisted-therapy-and-ptsd.pdf.

Itzchak, E. B., & Zachor, D. (2010). Who benefits from early intervention in autism spectrum disorders?. Research in Autism Spectrum Disorders, 5, 345-350. Doi: 10.1016/j.rasd.2010.04.018

Jasperson, R. A. (2010). Animal-Assisted Therapy with Female Inmates with Mental Illness: A Case Example From a Pilot Program. Journal of Offender Rehabilitation, 49, 417–433. Doi: 10.1080/10509674.2010.499056

Khanjani, Z. (2005). Development and pathology of interest from childhood to adolescence. Tabriz: Forouzsh Publications.

Komijani, Mehrnaz (2010). Childhood Anxiety Disorders. Exceptional Education, 100 & 101, 58-65.

Kruger K. A., Serpell J. A. (2010). Animal-assisted interventions in mental health: definitions and theoretical foundations, in Handbook on Animal-assisted Therapy: Theoretical Foundations and Guidelines for Practice, ed Fine A. H., editor. San Diego, CA: Academic Press, 33–48. Doi: 10.1016/B978-0-12-381453-1.10003-0

Last, C. G, Hansen, C., & Franco, N. (1998). Cognitive-behavioral treatment of school phobia. J Am Acad Child Adolesc Psychiatry, 37(4):404-11. Doi: 10.1097/00004583-199804000-00018

Malek, M., Hasanzadeh, R., Tirgari, A. (2013). Effectiveness of group play therapy and cognitive behavioral techniques in reducing behavioral problems in children with reading disorder. Journal of Learning Disabilities, 2(4), 140-153. doi: jld-2-4-92-5-8

Marguerite E. O., Noémie A. G & Alison C. K. (2015). Animal-Assisted Intervention for trauma: a systematic literature review. Front Psycho, 6, 1121. Doi: 10.3389/fpsyg.2015.01121

Mash, E. J., & Barkley, R. A. (Eds). (2014). Child psychopathology (3rd ed.). New York: Guilford Press.

Matin, A. (2009). Treatment of Anxiety Disorders in Childhood. Exceptional Education, 93 and 94, 32-32

McDowell, B. (2005). Nontraditional therapies for the PICU – Part 2. Journal for Specialists in Pediatric Nursing, 10, 81-85. Doi: 10.1111/j.1744-6155.2005.00009.x

Merikangas, K., Avenevoli, S. (2002). Epidemiology of mood and anxiety disorders in children and adolescents. In: Tsuang MT, Tohen M, editors. Textbook in Psychiatric Epidemiology. 2. New York, NY: Wiley, 657–704. Doi: 10.1002/0471234311.ch24

Monga, S., Birmaher, B., Chiappetta, L., Brent, D., Kaufman, J., Bridge, J., Cully, M. (2000). Screen for Child Anxiety-Related Emotional Disorders (SCARED): convergent and divergent validity. Depress Anxiety, 12(2):85-91. Doi: 10.1002/1520-6394(2000)12:2<85::AID-DA4>3.0.CO;2-2

Mychailyszvn, M. P., Mendez, J. L., & Kendall, P. C. (2010). School functioning in youth withand without anxiety disorders: Comparisons by diagnosis and comorbidity. SchoolPsychology Review, 39(1), 106-121.

Narimani, M., Soleymani, E., Abolghasemi, A. (2012). A comparison of internal and external dimensions of thinking styles in blind and sighted students. Journal of School Psychology, 1(1), 108-118. doi: d_1_1_91_3_1_7

Parish-Plass N. (2008). Animal-assisted therapy with children suffering from insecure attachment due to abuse and neglect: a method to lower the risk of intergenerational transmission of abuse?. Clin. Child Psychol. Psychiatry, 13, 7–31. Doi: 10.1177/1359104507086338

Prato-Previde, E., Custance, D. M., Spiezio, C., & Sabatini, F. (2003). Is the doghuman relationship an attachment bond? An observational study using Ainsworth’s strange situation. Behaviour, 140, 225–254. Doi: 10.1163/156853903321671514

Prothmann, A., Bienert, M., & Ettrich, C. (2006). Dogs in child psychotherapy: Effects on state of mind. Anthrozoos, 19, 265–277. Doi: 10.2752/089279306785415583

Rapee, RM., Kennedy, S., Ingram, M., et al. (2010). Altering the trajectory of anxiety in at-risk young children. Amer J Psychiat, 167(12), 1518-1525. Doi: 10.1176/appi.ajp.2010.09111619

Serpell, J. A. (2010). Animal assisted interventions in historical perspective. In A. H. Fine (Ed.), Handbook on animal‐assisted therapy: Theoretical foundation and guidance for practice (pp. 17–32). San Diego, CA: Academic Press. Doi: 10.1016/B978-0-12-381453-1.10002-9

Silverman, MJ. (2011). Effects of music therapy on psychiatric patients’ proactive coping skills: two pilot studies. The Arts in Psychotherapy, 38, 125-9. Doi: 10.1016/j.aip.2011.02.004

Southerland, E. M. (2007). A study of the effects of pet ownership on mental health among community-dwelling senior citizens in northeast Tennessee. Electronic Theses and Dissertations. Paper 2046. https://dc.etsu.edu/etd/2046

Tournier, I., Vives, M. F., & Postal, V. (2017). Animal‐assisted intervention in dementia. Swiss Journal of Psychology, 76, 51–58. doi:10.1024/1421-0185/a000191

Visagie, L., Loxton, H. & Silverman, W.K. (2015). Research Protocol: Development, implementation and evaluation of a cognitive behavioural therapy-based intervention programme for the management of anxiety symptoms in South African children with visual impairments. African Journal of Disability, 4(1), 160,10. doi:10.4102/ajod.v4i1.160

Worsman, M. (2016). animal Assisted Therapy for post traumatic stress Disorder (PTSD). Crockettfoudation.com

Yoosefi Looyeh, M., Delavar, A., Yoosefi Looyeh, M. (2008) The Impact of Narrative Therapy on Amelioration of Anxiety Disorder Symptoms in Forth Grade Students. JOEC, 8(3), 281-294.

Yount R., Ritchie E. C., St Laurent M., Chumley P., Olmert M. D. (2013). The role of service dog training in the treatment of combat-related PTSD. Psychiatr. Ann, 43, 292. Doi:10.3928/00485713-20130605-11.

